# Human-centred co-design of a dual-purpose heart failure dashboard

**DOI:** 10.64898/2026.09.20.26363523

**Authors:** Victoria Blake, Sarah Craig, Liesl Carvalho, Michelle Thomson, Jennifer Yu, Louisa Jorm, Nigel H. Lovell, Sze-Yuan Ooi, Blanca Gallego

## Abstract

**Introduction:** Heart failure care requires coordination across hospital and community settings, yet information is often fragmented across electronic medical records and clinical systems. Clinical dashboards can bring together and display information to support care delivery and service management; however, existing dashboards have generally focused on specific measures, interventions and monitoring pathways. This study aimed to co-design, iteratively develop and user-test an integrated heart failure dashboard that links patient-level clinical decision-making with service-level management.

**Methods:** A human-centred design approach comprising needs identification, collaborative ideation, iterative prototype development and end-user testing was applied across two complementary operational and clinical dashboard streams. Thirty-six clinicians, health service managers, data and implementation scientists, and consumers from metropolitan and regional services participated.

**Results:** For operational decision-making, participants prioritised real-time visibility of patients across heart failure services, patient trajectories, service-performance information, and identification of variation in guideline-directed care and outcomes. Clinical priorities included rapid synthesis of longitudinal information, optimisation of guideline-directed medical therapy, continuity across care settings, and clinical workload prioritisation. These requirements informed the dashboard prototypes. Many prioritised information elements were incompletely represented in structured data and distributed across disconnected systems and structured and unstructured clinical data sources.

**Conclusion:** Human-centred co-design identified complementary patient- and service-level information needs and translated them into linked dashboard prototypes. The proposed dashboard brings together current clinical status and longitudinal heart failure care history at the patient-level alongside service-level patterns. Implementation is required to evaluate whether these linked views improve care processes and patient outcomes.

## Introduction

Heart failure (HF) is a complex, chronic condition that imposes a significant burden on patients and healthcare systems worldwide, with rising prevalence driven by population ageing and increased cardiovascular risk factors.^1^ In Australia, approximately 30,000 new HF diagnoses occur annually, and HF remains the leading cause of hospitalisation in adults over 65, with many patients hospitalised at least once per year.^1,2^ HF-related admissions consume an estimated 1.4 million bed-days, and cost over one billion dollars annually.^2^ Importantly, up to one-third of readmissions are considered potentially avoidable, indicating significant opportunities to reduce system strain.^3^

Evidence-based interventions capable of reducing recurrent HF hospitalisation and improving quality of life are well established. These include optimisation of guideline-directed medical therapy (GDMT) during hospitalisation, cardiac rehabilitation and multidisciplinary HF disease-management programs, timely post-discharge review, and structured transitions between acute and primary care settings.^3^ Yet their implementation remains inconsistent. Australian data show incomplete prescribing of evidence-based therapies at discharge, low referral to cardiac rehab and HF home-visiting services, and low rates of post-discharge follow-up.^4,5^ This reflects both clinical constraints, such as comorbidity and advanced disease that may limit medication tolerability and service engagement, but also modifiable factors such as cautious prescribing, non-standard referral pathways, incomplete discharge communication, unclear ownership of post-discharge optimisation, limited access to HF services, low health literacy, limited delivery of disease-specific education, and fragmented care across hospital and community healthcare providers.^6,7^ Patient uptake of treatments and services is also suboptimal with poor HF specific health literacy, and personal and external barriers contributing to reduced engagement and adherence.^7^ These gaps disproportionately affect groups already facing higher HF burden including indigenous populations, people with lower socioeconomic status, culturally and linguistically diverse communities, and those living in rural and remote areas.^3,8,9^

Addressing these gaps may be supported by digital tools that provide timely visibility of potential care gaps and opportunities for care optimisation or additional support. However, clinical information relevant to identifying these gaps may be dispersed across multiple electronic medical record (EMR) systems and may be documented across structured fields and free text. These features can limit data completeness and make information synthesis inefficient and affect the ability of clinicians to make timely and informed decisions.^10^

Dashboards are visual information displays that integrate and present selected data in a structured format to support monitoring, interpretation and decision-making. By bringing relevant information into curated views, they can lower the cognitive burden on clinicians, make guideline-directed management easier to follow, identify care gaps at the point of care, support population-level monitoring of care quality, and enable more coordinated care across settings.^11–13^ However, their effectiveness depends on how well they align with end-user needs and fit the context in which they are used. A recent scoping review of 118 dashboards designed for diverse for healthcare settings and used for clinical and administrative purposes found that only half involved end users in the design process and 22% reported formative usability testing.^14^ Early and sustained engagement of end-users help identify relevant information and workflow requirements, while iterative prototype testing provides an opportunity to assess and refine how these requirements are translated into usable dashboard designs.^14^

Co-design and human-centred design approaches provide structured ways of involving end-users and other stakeholders in the design and development of clinical decision support (CDS) tools to ensure they are relevant, usable, and trustworthy.^15^ Co-design is defined as a collaborative process that brings together patients, caregivers and healthcare professionals to create technologies that are both clinically effective and user-friendly. Human-centred design strategies include scenario-based design, iterative usability testing, think aloud protocols, and explanation interfaces tailored to user needs, all of which improve interpretability and reduce cognitive burden.^16^ By embedding these methods within CDS development, tools are more likely to be adopted, integrated into workflows, and sustained in practice.

Existing HF dashboards have been developed for diverse purposes, including identifying patients for specialist review, monitoring readmissions and service performance, assessing guideline-concordant care, supporting GDMT optimisation and remote patient monitoring, and examining population-level monitoring of HF care and outcomes.^13,17–23^ Some provide links between aggregate indicators to patient lists or clinical profiles^13,22^, while other systems integrating clinical and administrative data across patient, service and organisational views.^19,24^ However, these systems have generally been organised around specific performance measures, interventions or care pathways. Less is known about the information and design requirements for a shared platform that supports both HF service management and patient-level clinical decision-making across hospital and community settings. Several systems have incorporated clinician feedback, Agile development and participatory design approaches.^19,25^ However, there is limited evidence describing multistakeholder co-design across clinical, operational and consumer perspectives across hospital and community settings.

### Objective

This study aimed to co-design an integrated HF dashboard supporting both patient-level clinical decision-making and service-level management across hospital and community-based care settings. Using a human-centred design approach, the project sought to identify current challenges and information, and workflow needs and translate them into a dashboard prototype with two complementary views:

1. **Operational view** – providing service-level insights into HF activity and outcomes to inform strategic planning and quality initiatives.
2. **Clinical view** – providing patient-level information to support bedside decision-making and facilitate shared decision-making between clinicians and patients.

## Methods

### Design Framework

The study design was informed by the human-centred design framework which encompasses five distinct phases or activities: empathise, define, ideate, prototype and test (Figure 1).^15^ This design process was repeated for both the operational and clinical views of the dashboard with the insights gained from the initial operational dashboard also informing the design of the clinical dashboard.

**Figure 1.**
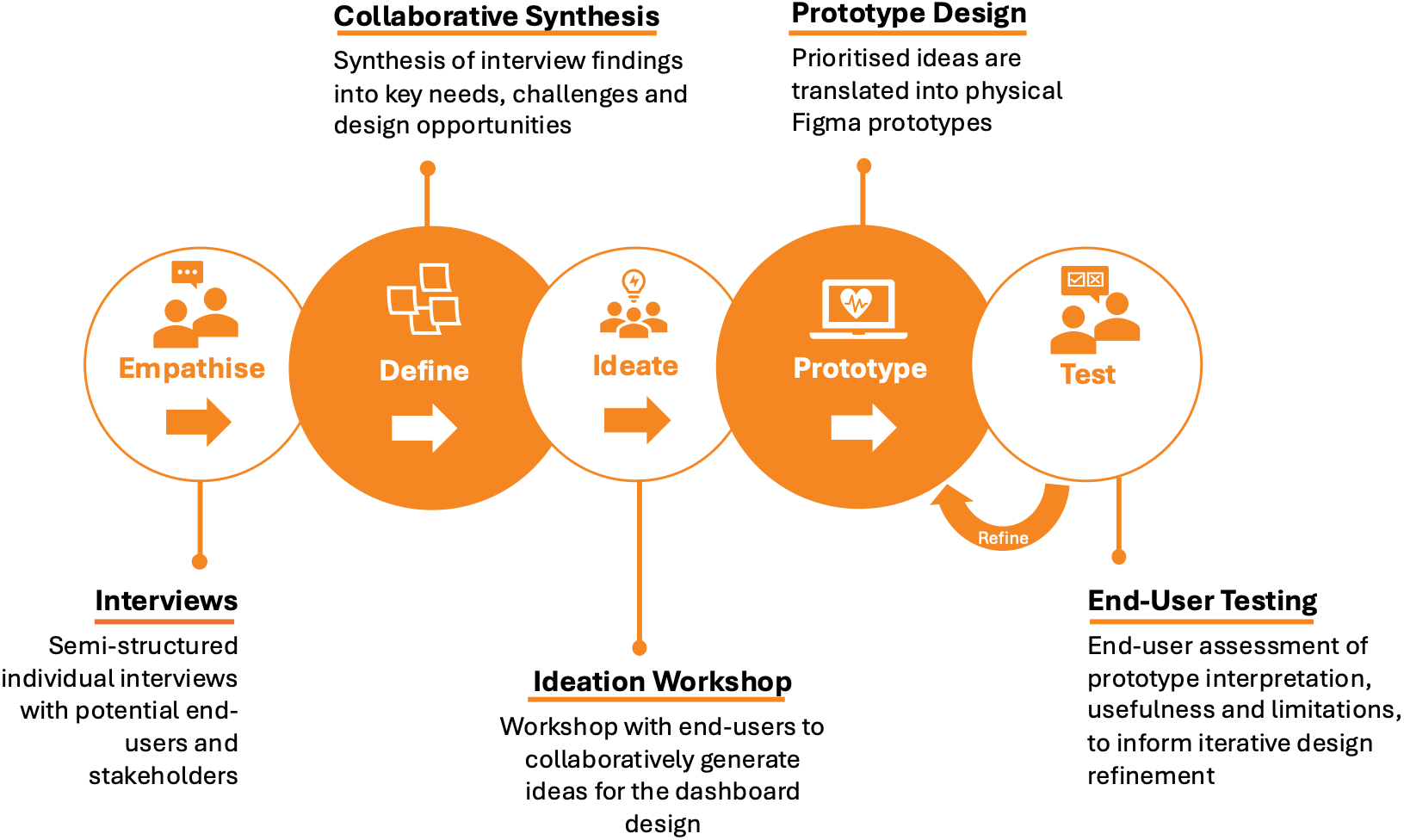
Human-centred design process used for development of the heart failure dashboard. The five-phase process comprised semi-structured interviews to understand end-user needs (empathise), collaborative synthesis to identify and define key needs and design opportunities (define), workshops to collaboratively generate and prioritise dashboard concepts (ideate), translation of prioritised concepts into physical and digital Figma prototypes (prototype), and end-user testing to assess prototype interpretation, usefulness and limitations (test). Testing findings informed iterative prototype refinement and retesting. The process was applied to both the operational and clinical dashboard streams, with prototype design and testing occurring iteratively. Adapted from the human-centred design framework described by Farao et al.^15^

### Setting

The study was conducted across two Local Health Districts (LHDs) in New South Wales (NSW), Australia, representing metropolitan and regional health service settings. South Eastern Sydney Local Health District (SESLHD) provides hospital and community healthcare to approximately 930,000 residents in metropolitan Sydney and includes major tertiary referral centre for cardiac services alongside public outpatient HF services. Mid North Coast Local Health District (MNCLHD) provides care to approximately 218,000 residents across a geographically dispersed regional area of northern NSW and provides interventional cardiac services but has more limited access to outpatient HF support.

### Participants and Recruitment

Participants were recruited from hospitals across both LHDs based on their roles and experience relevant to HF care, service delivery, digital health, dashboard development or lived experience of HF. Clinical and health service participants were initially identified through the professional networks of the clinical investigators, with snowball recruitment used to identify additional relevant stakeholders. Consumers with lived experience of HF were recruited through a community HF outreach service or via the eHealth New South Wales (eHealth NSW) Consumer, Community, Engagement & Partnerships Team.

The **operational dashboard stream** included hospital executives, cardiology heads of department, business intelligence staff, cardiac nurse unit managers, and digital health experts. The **clinical dashboard stream** involved cardiologists, junior doctors, community HF nurses, and consumers with lived experience of HF. All participants received an information sheet and consent form prior to participation, and verbal consent for either audio or video recording, or written documentation of the session, was obtained from each participant at the start of each interaction.

### Human-centred design process

#### Empathise

Semi-structured interviews were held with each participant individually and focused on understanding participant context, needs, challenges and opportunities in relation to HF patient care and service planning. Interview questions were adapted according to participant’s role and backgrounds. The semi-structured interview guide is provided in the Supplementary Material. Two investigators attended each interview, with sessions recorded using written notes, and where consented, audio or visual recordings.

#### Define

Following the interviews, the interviewers conducted a collaborative synthesis session using affinity mapping^26^, a human-centred design technique that supports the rapid identification of patterns and insights in qualitative data. Interviewers reviewed written notes and recordings, extracting key quotes and observations, transcribing them onto individual sticky notes. These were collectively organised into clusters representing user needs, challenges, and design opportunities, which informed subsequent ideation and prototype development.

#### Ideate

Separate one-day ideation workshops were held for the operational and clinical dashboard streams. Insights from the define phase were presented to workshop participants, alongside opportunity statements developed to guide idea generation. Structured activities were used to collaboratively generate, prioritise and design ideas that could be translated into prototypes and tested with end users. Workshop activities are described in Table 1. Outputs from the workshop activities were retained to directly inform subsequent prototype development.

**Table 1.**
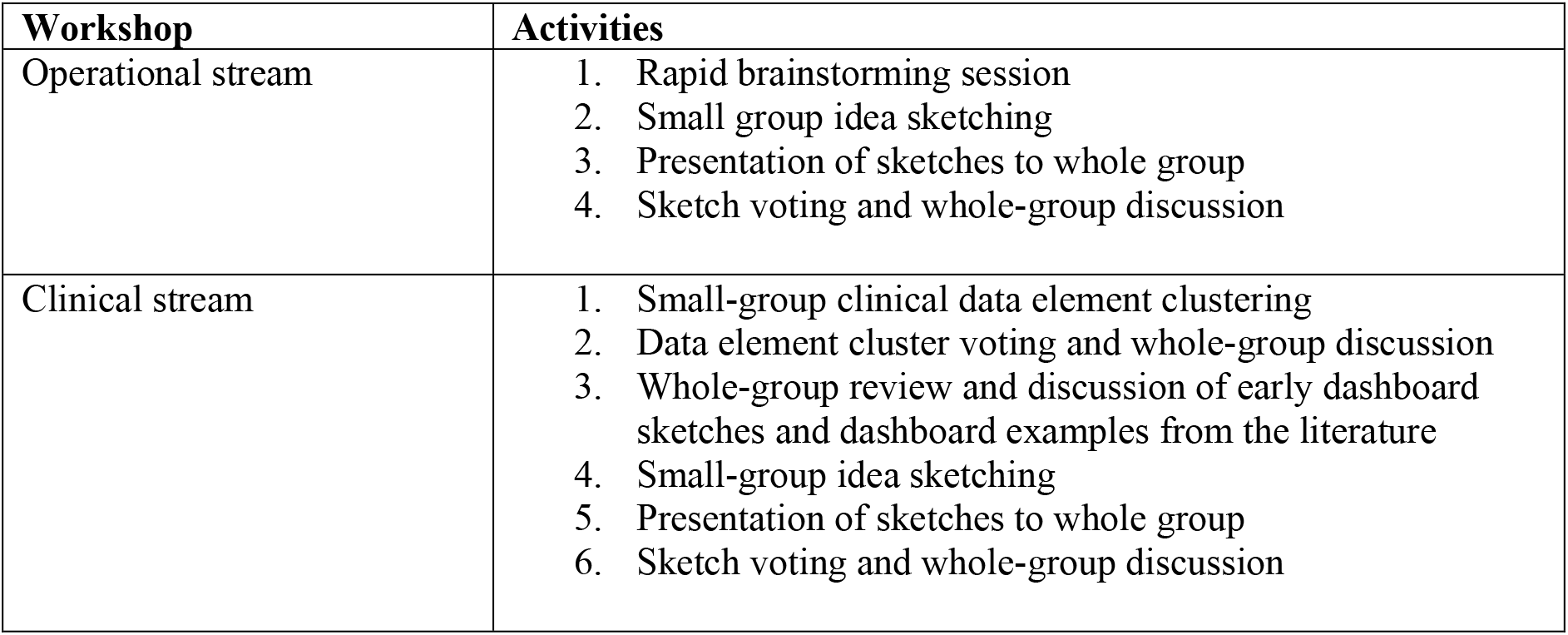
Ideation workshop activities.

| Workshop | Activities |
| --- | --- |
| Operational stream | 1. Rapid brainstorming session<br>2. Small group idea sketching<br>3. Presentation of sketches to whole group<br>4. Sketch voting and whole-group discussion |
| Clinical stream | 1. Small-group clinical data element clustering<br>2. Data element cluster voting and whole-group discussion<br>3. Whole-group review and discussion of early dashboard sketches and dashboard examples from the literature<br>4. Small-group idea sketching<br>5. Presentation of sketches to whole group<br>6. Sketch voting and whole-group discussion |

#### Prototype

Prioritised concepts and design ideas generated during the ideation workshops were translated into dashboard prototypes. Initial pen-and-paper sketches were progressively developed into interactive digital mock-ups using Figma (Figma, Inc.). Prototype content, information organisation and presentation were iteratively refined based on feedback from the end-user testing sessions.

#### Test

Formative end-user testing sessions were conducted in-person and virtually. Participants reviewed each prototype view without facilitator guidance and described their interpretation of the information presented before discussing its perceived usefulness, strengths, limitations, and opportunities for improvement. Feedback was reviewed by the design team and used to refine the prototypes, with revised designs subsequently retested with end-users.

#### Ethics

The project was reviewed by SESLHD Human Research Ethics Committee (HREC) who approved the project as a Quality Assurance/Quality Improvement (QA/QI) project.

## Results

### Participants

Across the two co-design streams, a total of 36 end-users and stakeholders participated in one or more co-design phases. Participants encompassed a range of roles including cardiology heads of department, senior and junior cardiologists, cardiac nursing unit managers, health system managers, health analysts and health data and implementation scientists, HF nurse practitioners, and consumers with lived experience of HF. Participants contributed to different phases of the co-design process, with some participating in multiple phases and across both dashboard streams. In total, 25 unique participants contributed to the empathise phases, 17 to the ideation, and 17 to prototype testing. Of the 36 participants, 17 contributed to more than one phase, including 7 who participated across all three phases of one or both streams. Figure 2 shows participation by role, phase and dashboard stream.

**Figure 2.**
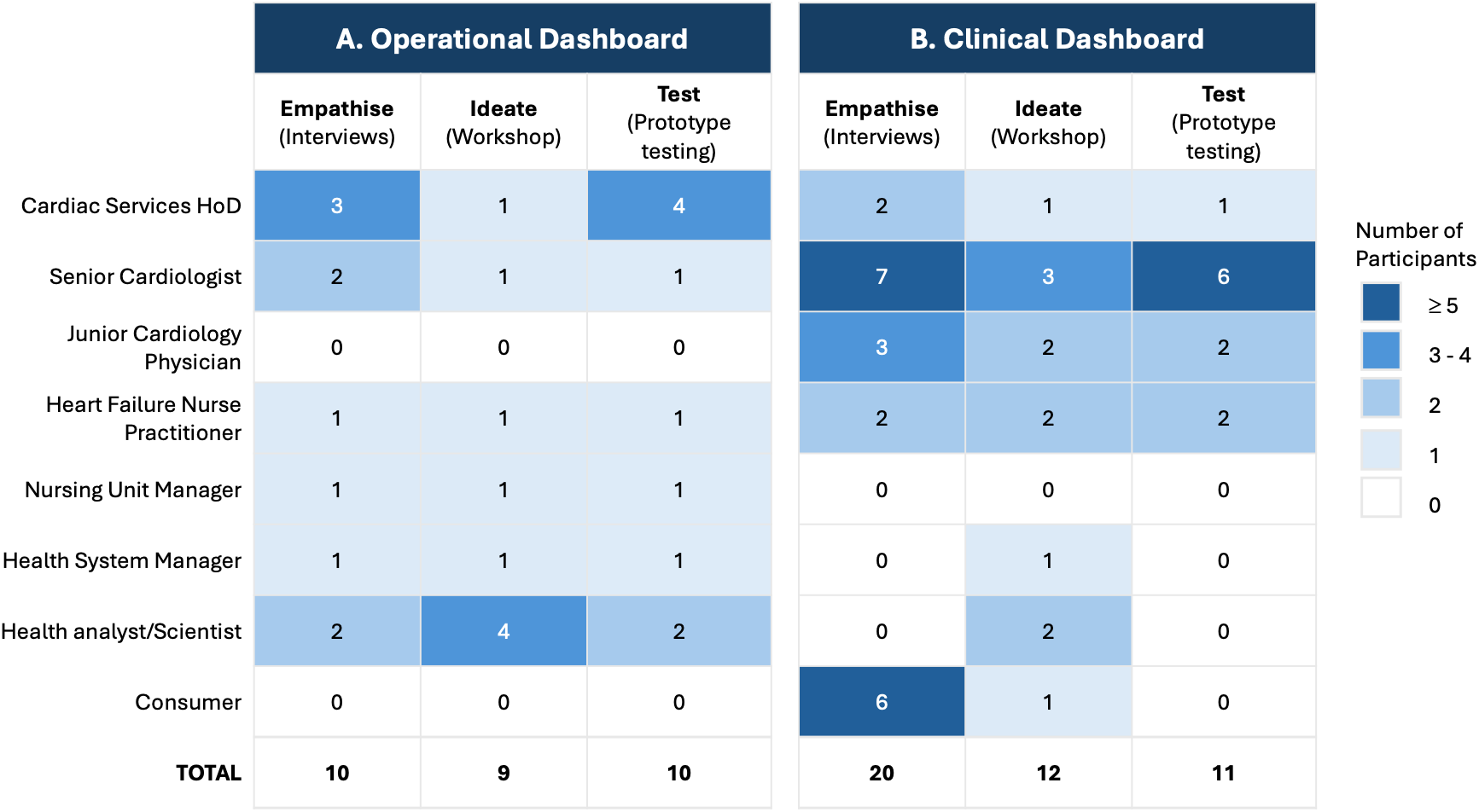
Number of participants by role and co-design phase for the operational (A) and clinical dashboard (B) streams. Numbers represent participants contributing to each phase. Some individuals participated in multiple phases and co-design streams.

### Operational Dashboard

#### Empathise and define

Participants described HF services operating under substantial time, workforce and bed pressures with clinicians managing patients across inpatient, outpatient and community settings. They reported limited access to timely service-level information, including which patients with HF were currently receiving care, where they were located and which hospital or community services they are connected with. This information often required manually searching the electronic medical record.

Participants wanted to identify how patients moved through the care network, identify gaps in treatment and referral, and understand whether variation in care and outcomes reflected differences in access to primary care, community HF services, rehabilitation or other resources.

Limited time and analytic capacity compounded this problem. One cardiologist described service data as *“not something at our fingertips every day”*, despite wanting to understand *“what works, what doesn’t work and where resourcing should go”*. Hospital managers similarly wanted high-level information on patient flow, readmissions, and service demand to guide resource allocation but admitted they lacked a HF specific lens. Key quotes related to these insights are presented in **Error! Reference source not found.**.

**Table 2.** Key operational dashboard needs identified during the empathise phase. Needs were identified through synthesis of semi-structured interviews with operational dashboard stakeholders. Illustrative participant quotations are presented for each identified need.

| Operational Needs | Key Quotes |
| --- | --- |
| Real-time visibility of patients across the HF service | *“We find it hard to find where all our patients are... a lot of our patients in a regional setting are admitted under general medicine”*<br>*“It’s not easy to go through 152 patients in EMR and see that somebody has been admitted somewhere in the health system”* |
| Understanding patient journeys across fragmented services | *“A lot of the challenge is understanding the big picture...having a longitudinal view of a patient’s journey...particularly in and out of other hospitals...would be really helpful for understanding where they are in their heart failure journey”*<br>*“Help people see there is a whole care network outside the hospital walls... we’re part of a journey, not the entire journey”*<br>*“We must ensure our patients get what they need no matter where they are”* |
| Making service performance data accessible and actionable | *“I’d like to understand in real time rather than after running a clinical trial to understand the effect of an intervention, a randomised control study can cost millions and take years”*<br>*“It’s less about the information not being available and it’s more about information being hard to find”*<br>*“I’d like to know how we perform compared to our peers of a similar size and across the state”*<br>*“There are challenges in understanding the service delivery... what works, what doesn’t work and understanding where resourcing should go”* |

#### Ideate, prototype and test

During the ideation workshop, participants generated and prioritised approaches for visualising the operational needs identified during the define phase, including patient location, patient journeys and service engagement, guideline-directed care and outcomes. Prioritised concepts informed the initial digital prototype, which were subsequently refined through three rounds of stakeholder testing. For the *“Where are our patients?”* view, testing led to progressive simplification of the initial design and greater emphasis on actionable patient groups (Figure 3). The refined view represented overlapping connections with community-based services and introduced filters to identify hospitalised patients already connected with the HF service and prioritise patients according to risk or need for review (not shown).

**Figure 3.**
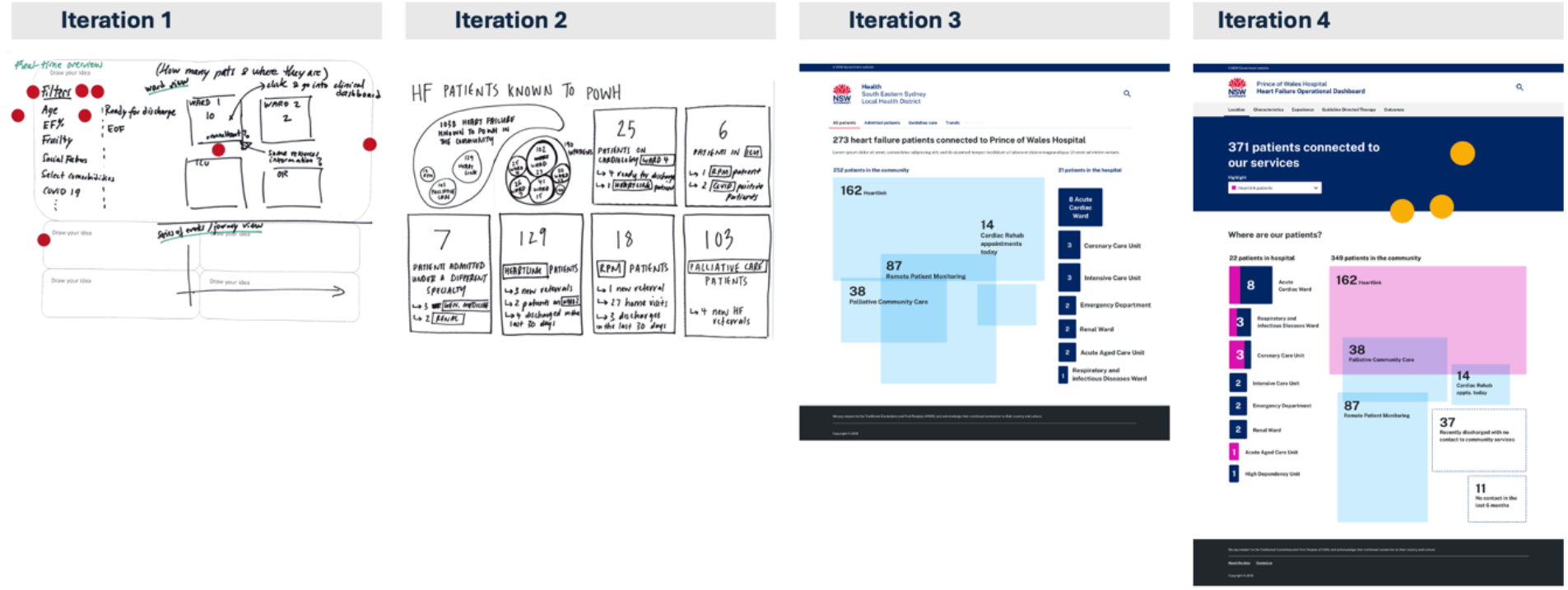
Iterative development of the operational dashboard “Where are our patients?” view. Iteration 1 shows concepts generated and prioritised during the ideation workshop. Subsequent prototypes were refined through rounds of end-user testing, resulting in a simplified visual overview, representation of overlapping community service connections, and functionality to identify and prioritise hospitalised patients for review.

Real-time visibility of patients across the HF service was addressed through the *“Where are our patients*?” view (Figure 4.A), which displayed the current distribution of patients across hospital wards and community services, including their connections with HF outreach, remote patient monitoring (RPM), cardiac rehabilitation and palliative care.

**Figure 4.**
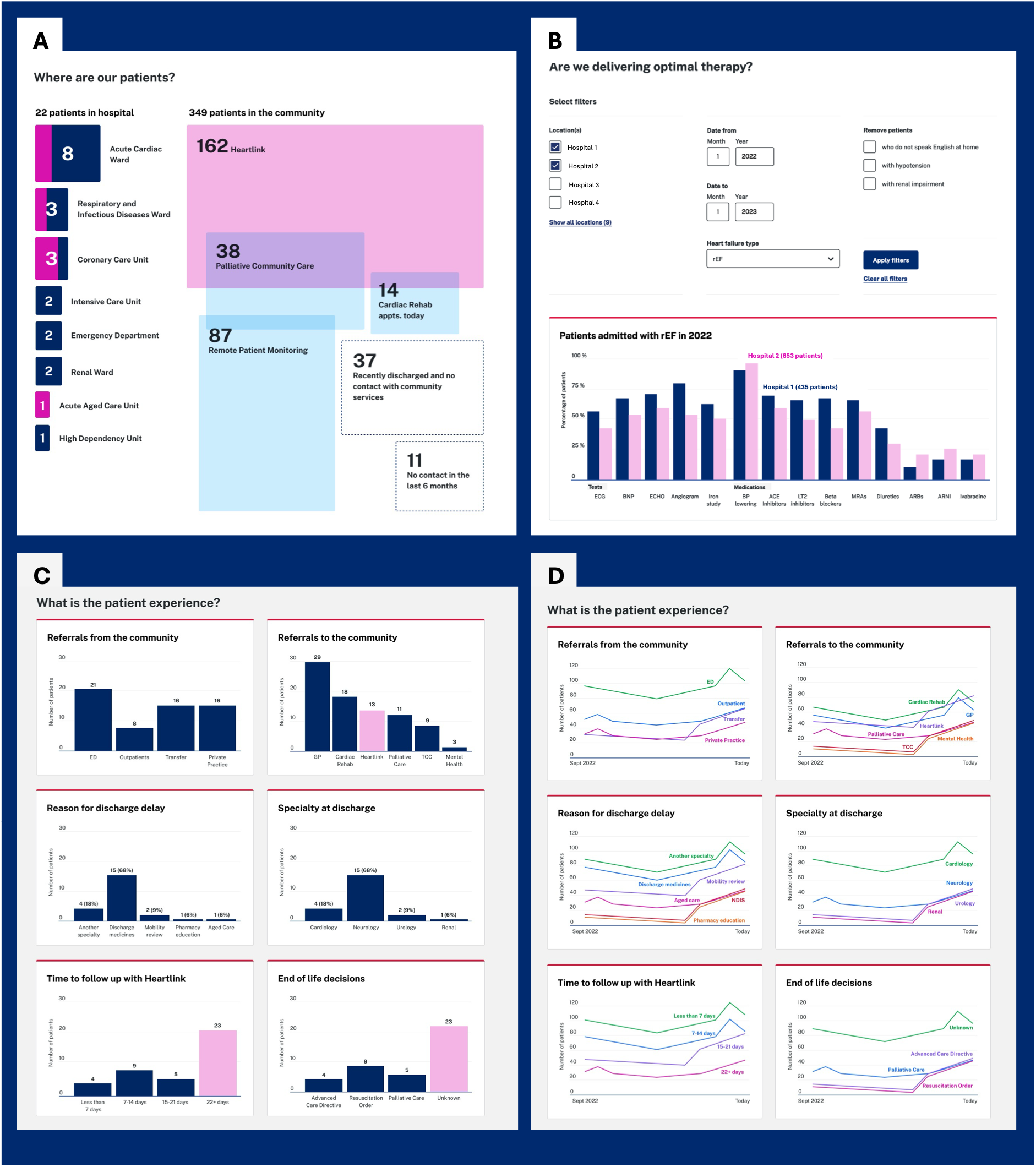
Selected views from the operational HF dashboard prototype. **(A) Where are our patients?** provides an overview of patients across hospital wards and community-based services. **(B) Are we delivering optimal therapy?** enables comparison of guideline-directed investigations and medical therapy between hospitals, with filtering by HF phenotype and relevant clinical factors **(C-D) What is the patient experience?** presents referral pathways, discharge delays, speciality at discharge, time to HF service follow-up and end-of-life planning as aggregate comparisons **(C)** and longitudinal trends **(D)**

Making service performance data accessible and actionable informed several complementary views. The *“Who are our patients?”* view characterised the HF population by demographic and clinical factors, while the *“Are our patients receiving optimal therapy*?” view (Figure 4.B) allowed users to examine delivery of recommended investigations and GDMT and compare performance between sites. The *“What are our outcomes?”* view (not shown) summarised length of stay, readmissions, and mortality.

Understanding patient journeys across fragmented services was less comprehensively addressed through the co-design process. The “*Patient Experience*” view (Figure 4.C & D) incorporated measures of referrals, discharge delays, time to community HF follow-up and end-of-life planning; however, these aggregate indicators did not fully represent the longitudinal movement of patients through the care network identified as important during co-design. Further work was therefore planned to develop and evaluate approaches for visualising end-to-end patient pathways.

Participants particularly valued the *“Where are our patients?”* view because it provided clear, clinically meaningful information not currently available that could directly support day-to-day operational efficiency. One participant noted that identifying patients before ward rounds could *“free (the HF nurse) up to go see people in the community as opposed to hanging around for two hours”*.

The *“Are our patients receiving optimal therapy?”* view also received positive feedback. Participants valued the ability to assess adherence to guideline-directed care, understand variation and compare sites, considering this useful for quality improvement and resource planning. One participant explained *“This is what we want to see. Are we being compliant with guideline things and if we’re not why?”.* Participants also emphasised the importance of documenting reasons for deviation from GDMT, allowing patient-level clinical context to inform interpretation of aggregate measures.

The *“Who are our patients?”* and *“Patent experience”* views received mixed feedback. Although considered potentially useful, some questioned whether the aggregate charts adequately explained the patterns shown, with one participant asking, *“what is this data actually telling me?”*.

Across the views, participants emphasised the importance of linking operational measures to patient-level clinical information so that service-level variation or care gaps could be investigated and acted on at the bedside. Participants felt the two views would be accessed for different primary users and workflows rather than as a single interface used by all participants. However, the proposed linkage would allow most users to access the *“Where are our patients?”* view with authorised users drilling down into the patient-level clinical view or moving through the service-level views in the operational dashboard.

### Clinician Dashboard

#### Empathise and define

Clinicians consistently described difficulty accessing and synthesising longitudinal patient information across clinical notes, investigations, medication records and other parts of the EMR (Table 3). Reconstructing patient histories was time-consuming, dependent on clinicians’ information seeking skills, and could delay decision-making or lead to repeat investigations. One participant stated that relevant information *“should be at the touch of a button, so clinicians don’t spend 30-60 minutes gathering information on a patient*.

**Table 3.** Key clinical dashboard needs identified from stakeholder interviews during the empathise phase. Needs were identified through collaborative synthesis of interview findings. Illustrative participant quotations are provided to demonstrate the perspectives relative to each need.

| Clinical Needs | Key Quotes |
| --- | --- |
| Difficulty accessing and synthesising complex patient histories across fragmented systems | *“I think clinically a lot of the challenge is understanding the big, picture...[having] all the information you need to make an appropriate assessment...having a longitudinal view about that patient’s journey, particularly if their journey takes them in and out of other hospitals.”*<br>*“Residents spend a large part of their job tracking down results... we don’t want to repeat invasive or costly tests.”*<br>*“When I’m on call the quality of the information I get depends on how good the ED intern is at finding it in the EMR”*<br>*“There’ll be a day or two trying to gather correspondence before we start investigating things... so as we don’t replicate investigations.”* |
| Difficulty identifying and acting on opportunities to optimise HF care | *“You see someone who’s not on something and you’re like, oh, that’s odd. Why they’re not on that drug... kidney function went bad... blood pressure went low... heart rate went low.”*<br>*“Every three months we have a new set of Junior Medical Officers and our delivery of therapy varies due to that... it can delay treatment and increase length of stay.”*<br>*“We could have started a more aggressive treatment earlier... we’re now up to 10 days admission because we waited.”*<br>*“We need to give patients interventions before they start to deteriorate”* |
| The need to prioritise clinical workload | *““If you got someone with a 62% readmission rate... forget about the other people... go and see this one. So there’s a batting order of people to see.”*<br>*“I want to see only my patients, prioritised by who I should see first... who is sick, who is waiting to be discharged.”* |
| Poor continuity across hospital, specialist, primary care and community settings | *“Discharge letters are suboptimal, sometimes without a clear plan for the GP or specialist... If can’t decipher it, I’m not sure how the patient is going to”*<br>*“We need to make end of life information available to us it can take us 24 hours to find out a patient’s preference was to be kept comfortable”*<br>*“It’s a minefield, you can’t be sure what (medication) a patient is taking... if there’s a mistake made it carries through the whole admission”*<br>*“Once they leave hospital it’s up to the GP to follow our plan for blood tests and med changes, which doesn’t always happen”* |

Fragmentation also made opportunities to optimise HF care difficult to identify. Clinicians wanted ventricular function, medication initiation and titration, renal function, adverse effects and contraindications to be brought together so that potential gaps in guideline-directed care could be distinguished from clinically appropriate treatment decisions. Poor information continuity across hospital, specialist, primary care and community settings compounded these problems, particularly for medication changes, discharge plans and investigations performed elsewhere.

Clinicians also wanted the dashboard to support prioritisation of limited clinical resources. Patient lists that identified deterioration, prolonged admission or discharge readiness were considered particularly useful for HF outreach services managing large community caseloads and hospital teams caring for patients dispersed across different wards or regional hospitals.

Consumer perspectives (Table 4) reinforced that fragmentation was experienced not only as an information problem for clinicians, but as a continuity-of-care problem for patients. Consumers described repeatedly recounting their histories, variable communication between hospital and primary car and both patient and clinician uncertainty about medications. Some reported limited understanding of their condition and self-management after discharge, while others maintained their own medication lists to support communication. Consumers expressed interest in accessing the dashboard to support conversations with clinicians and verify their medical information and reported limited privacy concerns provided its use was considered safe and improved care.

**Table 4.** Insights from consumers with lived experience of heart failure.

| **Consumer Perspectives** | **Key Quotes** |
| --- | --- |
| Consumers are having to repeatedly recount their medical histories | *“When I get to hospital, they ask me all these questions I can't answer”*<br>*“You have to repeat yourself and repeat again”* |
| Consumers highly value the care received through the outpatient HF-management service | *“They are very good... the nurse is just running around the whole world... but whenever you call, they pick up”*<br>*“I could always call the nurse if I needed... It's reassuring to know they are there if I need them”* |
| Consumers described a lack of coordinated care across settings | *“I didn't have a regular GP to prescribe what I needed*<br>*“In the hospital they take care of everything ... in the real world the GP said they didn't know about heart failure”*<br>*“The GP didn't know what my medicines were for”* |

#### Ideate, prototype and test

During the ideation workshop, participants reviewed the needs identified during interviews and grouped and prioritised clinical information according to how it should be organised for patient care. Small multidisciplinary groups then developed dashboard concepts, while non-clinical participants considered how consumer insights could inform a patient-facing view. Prioritised sketches and workshop decisions informed development of the clinical prototype.

The prototype brought together related information around the patient’s HF trajectory. The *“Current summary”* view (Figure 5) provided an initial overview of HF history and diagnosis, current GDMT, recent physiological trends, admissions and investigations. The prototype also included illustrative displays of predicted readmission and mortality risk to explore how information from a proposed AI model could be incorporated into the dashboard. Drill-down views provided longitudinal detail, including an *“Admission history”* timeline and *“Medications at discharge”* view showing GDMT initiation, dose changes and treatment duration. The *“AI-generated risk at discharge”* view displayed risk trajectories and variables contributing to predicted risk.

**Figure 5.**
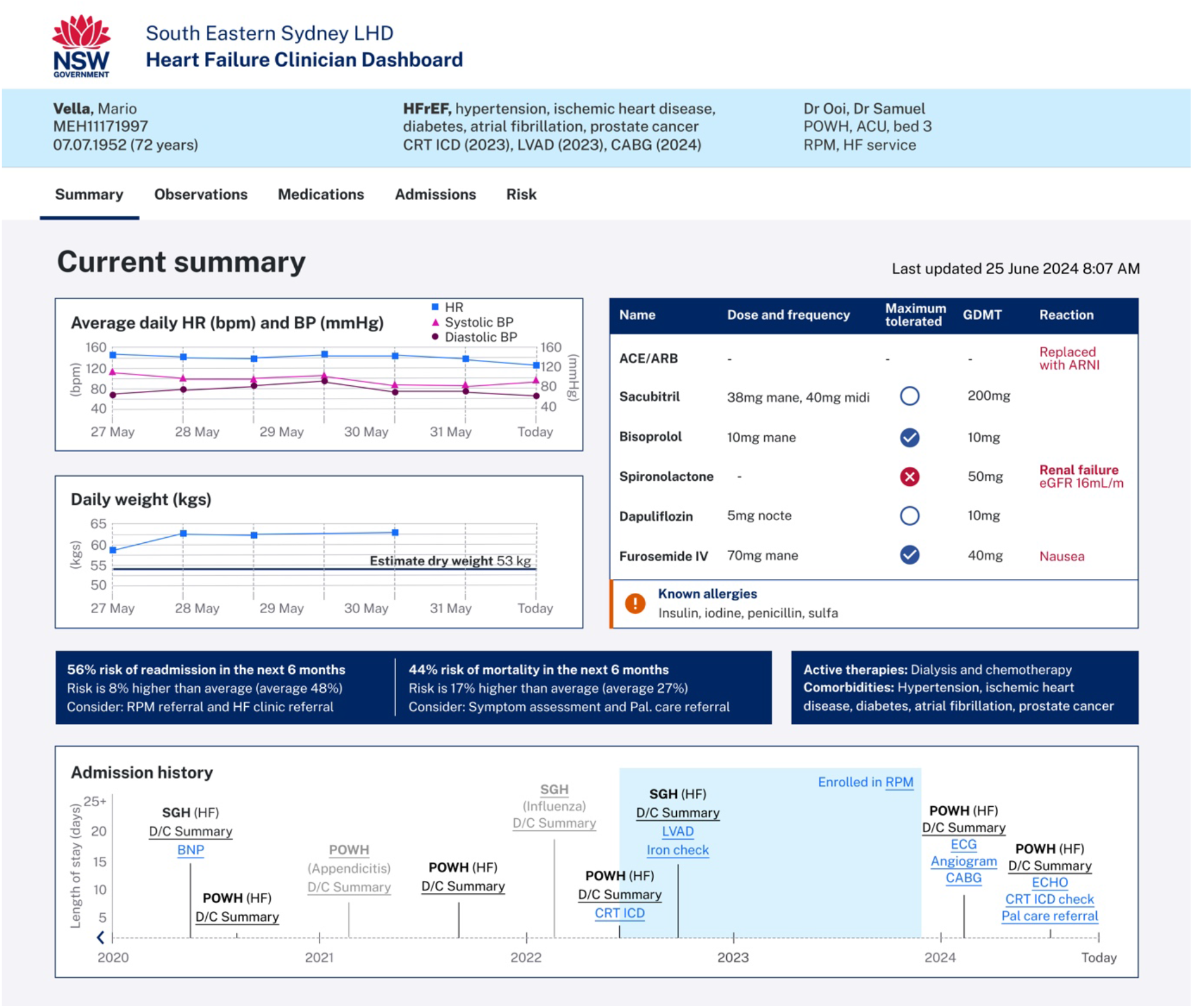
Clinical dashboard “Current summary” view. The summary-first view brings together current physiological status, guideline-directed medical therapy and treatment limitations, predicted risk, active therapies and comorbidities, and longitudinal admission and care history, with links to underlying clinical information for drill-doen review. All patient information and risk estimates shown are illustrative and do not represent real patient data or outputs.

Participants particularly valued the *“Current summary”*, *“Admission history”* and *“Medications at discharge”* views because they reduced the work required to reconstruct the longitudinal record. The patient summary was described as *“a nice snapshot”,* supporting rapid orientation without requiring multiple clicks, particularly during busy ward rounds with unfamiliar patients with a participant noting *“It’s good because it’s there all in the one page… I’d have to scroll through EMR and go back through multiple admissions to get all that information.”*. Testing informed further refinement to add a banner to make HF phenotype, relevant comorbidities, HF service involvement and active therapies immediately visible, and expansion of the timeline to include all encounter types with direct links to source documents, allowing the dashboard to function as an entry point to the underlying clinical record rather than replacing it. Subsequent feedback identified potential information overload leading to additional filters, hover summaries and functionality to focus on selected time periods.

The longitudinal *“Medications at discharge”* (Figure 6.A) view received consistent positive feedback. Participants valued being able to see the pillars of GDMT, dose changes, treatment duration and reasons for discontinuation or dose reduction, with one describing medications as *“much more visible”* than the existing EMR. Participants emphasised that medication changes made outside hospital, including by community HF services, were also needed. Linking treatment changes with renal function, blood pressure and other clinical parameters was considered important for providing context for apparent gaps and distinguishing opportunities for initiation and uptitration from previous intolerance, adverse effects or other clinically appropriate reasons for deviation from GDMT. In response, functionality was added for users to input reasons for GDMT deviation.

**Figure 6.**
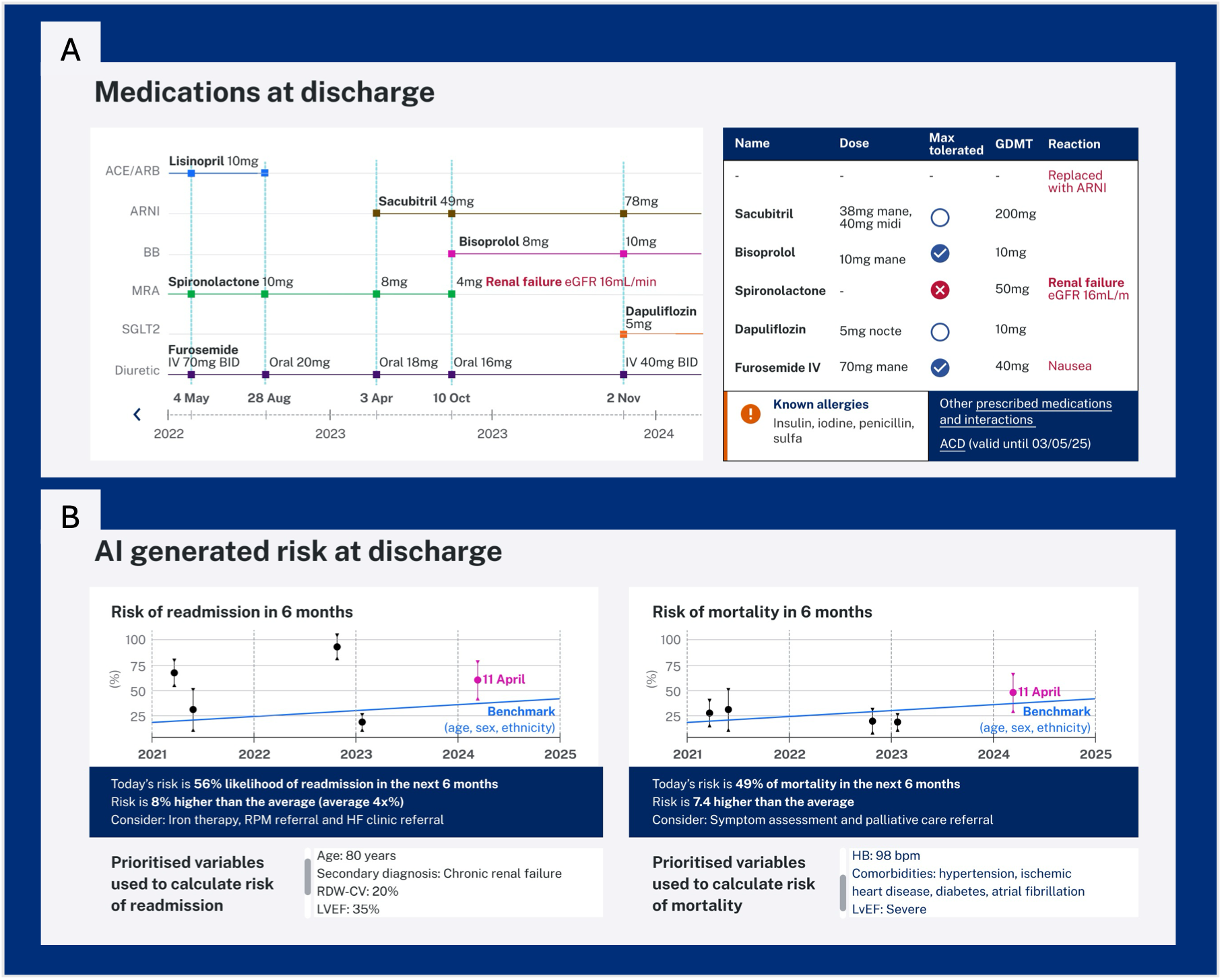
Clinical dashboard prototype. **(A) “Medications at discharge”** view showing current and longitudinal prescription and dosage of HF GDMT drug classes for an individual patient; and **(B) “AI-generated risk at discharge”** view showing the proposed presentation of current and longitudinal predicted readmission and mortality risk and factors contributing to those predictions. All patient information and risk estimates shown are illustrative and do not represent real patient data or outputs.

Feedback on the proposed presentation of predicted readmission and mortality risk in the *“AI-generated risk at discharge”* view (Figure 6.B) was mixed. Some participants considered predicted risk potentially useful for prioritising limited HF resources, prompting palliative care referral, and supporting prognosis conversations. Initial prototypes presented predictions using low-, medium- and high-risk categories, but participants questioned how these thresholds would be defined. In response, categorical risk bands were replaced with continuous risk estimates presented as trends and relative to other patients with HF, with confidence intervals. Others questioned which underlying variables would underpin the predictions and whether clinical and social factors would be adequately represented, with one participant stating, *“I’d need to know what’s behind it”.* Participants emphasised the need for transparency about model inputs, limitations and uncertainty if risk estimates were to support clinical assessment.

Some cardiologists cautioned that the HF-focused design could obscure clinically important non-HF context or encourage an overly formulaic approach to guideline-directed care. One highlighted comorbidities and treatments that may precipitate decompensation or constrain HF management, such as cancer therapy or corticosteroids, and the need to retain a broader clinical context without creating information overload. Others worried that presenting GDMT as a checklist could inadvertently encourage clinicians to prioritise guideline targets over individualised care and patient priorities.

#### Overall design and implementation considerations

Across both dashboard views, participants emphasised that usefulness would depend on providing timely, trustworthy information within existing workflows. Iterative testing identified that participants consistently preferred a summary-first, drill-down design, with key information available for rapid orientation and more detailed longitudinal views or source documents accessible when required. Minimising navigation and loading time were considered particularly important during ward rounds, where limited hardware, a slow network, and multiple EMR clicks already disrupted workflow. Clinicians also highlighted the value of mobile or tablet access. Table 5 provides insights around implementation from participants.

**Table 5.** Dashboard Implementation considerations identified through co-design. Illustrative participant quotations highlight usability and workflow requirements and the importance of data trustworthiness for future implementation.

| **Insights** | **Key Quotes** |
| --- | --- |
| Usability and workflow integration concerns<br>(availability of hardware, need for mobile/tablet access) | *“The screens are too small to view a dashboard and make entries into the EMR and view results.”*<br>*“A tablet would be great for people looking at information and making decisions”*<br>*“The great thing about paper is the fact that you can flick through a large amount of information quickly.* |
| The importance of trust in the data | *“If we can demonstrate the evidence behind what we’ve built, that builds trust”*<br>*“Make it clear there are gaps...omission of details is important. If you think you have the most up to date tests and you don’t, it’s a problem”* |

Participants also emphasised the importance of data provenance, recency and completeness. Integrating information across fragmented systems was considered a major potential benefit of the dashboards, but stakeholders questioned the feasibility of maintaining accurate and timely data across multiple hospitals and services. Missing or unvalidated information needed to be clearly identified, with access to underlying source documents where appropriate. This was particularly important for information derived from clinical text or predictive models, where clinicians wanted to understand the source and limitations of the information presented.

Finally, participants stressed that aggregate measures required appropriate clinical and service context. Variation in GDMT use could reflect contraindications or intolerance rather than a gap in care, while differences in service outcomes could reflect patient populations, workforce or resource availability. Participants cautioned that operational monitoring and benchmarking could undermine engagement if used punitively rather than to identify opportunities for improvement. These findings highlighted the need for implementation and governance approaches that preserved clinical context and supported quality improvement while ensuring the dashboards remained reliable, transparent and aligned with clinical workflow.

#### Data and technical requirements

Meeting the information needs identified through co-design would require integration of structured and unstructured data across multiple hospital and community systems. Required information included demographics, service engagement and outcomes at the operational level, and longitudinal medications, investigations, physiological trends, comorbidities, functional status and care transitions at the clinical level. Relevant data were distributed across the hospital EMR, pathology and imaging systems, pharmacy records and other clinical databases.

Many high-priority elements are predominantly documented in unstructured clinical text (Table 6). HF phenotype, functional and symptom status, reasons for deviation from GDMT, and discharge planning were predominately documented in clinical text, while information such as echocardiogram findings could be distributed across structured measurements, reports and external systems. Natural language processing and integration of existing structured data will therefore be required to provide the longitudinal clinical context prioritised by participants.

**Table 6.** Information to be sourced from unstructured data for the dashboard.

| **Information Type** | **Data Elements** | **Format within the EMR** |
| --- | --- | --- |
| HF diagnosis and phenotype | Mentions of HF diagnosis<br>Phenotypes such as HFrEF, HFpEF, Left HF, Right HF | ICD codes provide HF diagnosis, but they are only coded after discharge, and HF phenotypes are not currently included in ICD coding systems |
| Functional status and symptom burden | NYHA score<br>Oedema,<br>Dyspnoea,<br>Chest pain,<br>Exercise tolerance<br>Independence with ADLs | All symptom and functional information exist within clinical notes only |
| Echocardiogram results | Left Ventricular Ejection Fraction (LVEF),<br>Structural heart abnormalities,<br>Valve regurgitation and stenosis | Echocardiogram results are recorded in a mix of structured measurement, images and text reports, however these are often kept in separate, unlinked echocardiogram database.<br>The text reports are copy pasted into EMR clinical notes.<br>Outpatient echo reports scanned as pdfs into EMR or transcribed into clinical notes. |
| End-of-life planning and care | Advanced care directives, palliative consultations, discussions with patients and families about the goals of care. | Some end-of-life planning documentation exist within structured forms.<br>Many end-of-life planning details contained within the clinical notes. |
| Comorbidities & reasons for GDMT deviation | chronic kidney disease, hypotension, chronic obstructive pulmonary disease, atrial fibrillation, ischaemic heart disease diabetes, anaemia and iron deficiency, frailty and cognitive decline, depression and anxiety, liver disease, malignancy | Some items are contained within diagnosis codes or structured data.<br>Decisions around GDMT in relation to patient's comorbidities are only documented in the clinical notes. |

## Discussion

In this study, we used a human-centred design process to identify user needs, develop and iteratively test linked operational and clinical HF dashboard prototypes. User testing refined how longitudinal clinical and service information should be organised and surfaced several requirements for implementation. Visibility of patients and care events across settings, alongside information about GDMT, current clinical status, service engagement and risk were prioritised by participants. These priorities align with established challenges in HF care, including inconsistent optimisation of evidence-based therapy and fragmented transitions between hospital and community care.^3,4^ Participants described a central barrier to improvement as the difficulty of accessing, integrating and interpreting existing information in a way that supports day-to-day decision-making. The dual-view design translated these priorities into a prototype intended to support identification and investigation of potential care gaps at both service and patient levels. This co-design project documents the perceived usefulness of the proposed tool and provides data and system integration specifications for future implementation.

Consumer accounts indicated that fragmented systems transfer part of the burden of maintaining continuity onto patients, who repeatedly recount their medical history, reconcile medication changes or maintain their own records. The potential value of an integrated dashboard may extend beyond clinician efficiency to support continuity, shared understanding and patient participation. However, how patients should access and interact with dashboard information will require further co-design, particularly around appropriate content, and processes for correcting inaccurate information.

The principal design contribution of this work is the integration of service-level HF management with a longitudinal clinical view intended to support patient-level decision-making within a shared platform. Although previous systems allow movement from aggregate indicators to patient lists or profiles, they generally have been organised around specific performance measures, interventions or monitoring pathways.^13,20–22^ Our operational view encompasses patient flows, service engagement and outcomes, while the clinical view brings together longitudinal histories, comorbidities, GDMT, and risk assessment.

Stakeholders wanted these levels to be connected where relevant, for example, using population-level variation in GDMT, or risk to identify priorities and unmet needs, then drilling down to individual patients and contextual factors underlying that variation and risk, and the appropriateness of potential interventions.

Identifying a potential care gap and determining whether it is actionable are therefore distinct information tasks. Studies using the Veterans Affairs (VA) HF dashboard demonstrate this distinction. The randomised DASH-HF trial evaluated whether dashboard-directed telehealth clinics could improve the use and dosage of GDMT among patients with HFrEF that were identified as having opportunities for treatment optimisation, but the trial did not significantly improve its primary measure of GDMT optimisation.^23^ A subsequent implementation of the same dashboard with a population health clinic similarly demonstrated the importance of additional context to explain GDMT gaps, finding 44.8% of patients identified by the dashboard as having GDMT deficiency had at least one apparent treatment gap that was considered clinically inappropriate after clinical review, commonly because of previous intolerance, phenotype misclassification, incomplete medication capture, hospice care or renal impairment^27^. These successive implementation studies reinforce our findings on the importance of capturing the treatment rationale, broader clinical context, underlying information and data provenance. They also align with participants’ concern that prominent guideline-based indicators could encourage formulaic care or obscure clinically appropriate variation. Variation from guideline-directed care is not necessarily unwarranted and may reflect clinical complexity, treatment tolerability, resource availability, or patient needs and preferences.^28^ The clinical view must therefore preserve access to this context without increasing information overload.

A further contribution of this work is the translation of user needs into an informatics specification. Many information elements prioritised by participants are incompletely represented in structured EMR fields and instead occur within clinical notes or siloed clinical systems. Delivering the proposed dashboard therefore requires not only visualisation, but an underlying data infrastructure capable of integrating longitudinal information across these sources and extracting clinically meaningful concepts from unstructured text. Previous HF dashboard implementations illustrate this requirement. The VA HF dashboard combined structured data with NLP-derived ejection fraction and HF phenotype from unstructured clinical and imaging reports, with subsequent optimisation incorporating data-source hierarchies to improve HF phenotype classification.^21,22^ However, these implementations also demonstrate that inaccurate phenotype classification, incomplete medication capture and ambiguous source data can affect dashboard outputs, requiring clinical validation and transparent data provenance.^21,22,27^ The data infrastructure to support our dashboard implementation is being developed through the CardiacAI project,^29^ which brings together EMR and longitudinal outcomes data across participating health services alongside natural language processing methods to derive information that cannot be reliably obtained from structured data alone.

However, completeness will remain constrained where clinically important events occur outside of connected systems. Medication initiating or dose changes made in primary care, private specialist services or other health services may not be visible, or only partially, making longitudinal treatment trajectories and care gaps difficult.^21,27^ Even with improved integration, transparently presenting information that is absent, outdated, unavailable from another system or not identified by automated extraction will require careful validation and representation of uncertainty, provenance and recency.

### Limitations and Future Work

Despite the strengths of a multi-stakeholder, iterative process, several limitations should be acknowledged. Most clinician participants were drawn from a single health service, which may have constrained the diversity of perspectives and limited the generalisability of specific design preferences to other settings. Power dynamics in workshop settings, particularly where senior clinicians and managers were present, may also have influenced the willingness of some participants to voice disagreement or propose more radical ideas, a well-recognised limitation of user- and human-centred design in healthcare^30^. The inclusion of multiple professional groups and consumer representatives, separate operational and clinical streams, and one-on-one interviews and prototype testing sessions helped to mitigate some of these risks.

Prototype testing was formative and focused on ease of interpretability, perceived usefulness and design refinement rather than formal usability metrics and task performance. Evaluation of usability, workflow integration and effects on clinical decision-making will therefore need to be undertaken during implementation. Some requirements identified through co-design, particularly timely integration of information across systems and transparent representation of data completeness, will require technical development and evaluation in the live environment. The prototypes specify information and navigation requirements, but whether they improve clinical decision-making or action remains to be evaluated.

The next phase of this work will develop and trial these prototypes as a live dashboard through a funded implementation program, with further co-design focused on several components. First, the operational dashboard prototype did not yet fully represent the patient trajectories in relation to treatment, service connections and outcomes, and the workflows for navigating between operational and patient-level views require further testing. Further work will explore approaches for representing these relationships. Second, development of the predictive risk score has commenced,^31^ but will require additional co-design and silent evaluation of usability and acceptability including how explainability and uncertainty can be presented in ways that support rather than displace clinical judgement. Finally, implementation will allow evaluation of whether integrated clinical and operational views improve care processes, including identification of treatment and service gaps, coordination of care, service utilisation and patient outcomes.

## Conclusion

This study identified the information, navigation and integration requirements for a shared HF dashboard linking service-level management with patient-level clinical decision-making. Participants valued the ability to rapidly understand a patient’s current status and longitudinal history of HF care within an integrated dashboard that bridges operational and clinical perspectives. They also emphasised that the information must be reliable, support contextual interpretation and align with existing workflows. This co-design process has generated a blueprint for future implementation and evaluation. Subsequent work will need to translate these prototypes into live systems, develop the necessary data integration and text extraction pipelines, and rigorously assess their effects on care processes, delivery of guideline-directed care, and patient outcomes.

## Funding

This work was supported by the Australian Government Medical Research Future Fund Cardiovascular Health Mission (grant number [2021/MRF2008991]).

## Acknowledgements

The authors thank the clinicians, health service managers, data and implementation specialists, and consumer representatives from SESLHD, MNCLHD, and NSW Health who generously contributed their time, experiences and perspectives throughout the co-design process. We gratefully acknowledge eHealth NSW for providing human-centred design expertise, facilitating co-design activities and developing the Figma prototypes for this project.

The authors used OpenAI ChatGPT to assist with language editing, manuscript refinement and improving the clarity and structure of the manuscript. All AI-assisted content was reviewed and revised by authors, who take full responsibility for the final manuscript.

## Data Availability

Interview recordings, notes and other co-design materials generated during the co-design process, and not presented here or in the supplementary material, are not publicly available because they contain potentially identifying participant information and were collected under consent and governance arrangements that did not provide for public data sharing.

## Disclosures

The authors declare no competing interests.

## Supplementary

### 1. Operational dashboard stream – Stakeholder interview conversation guide

| Key question | Ask these as prompts if needed |
| --- | --- |
| Tell me a bit about **you and your role**<br>(5 mins) | How long have you worked at _____?<br>What is your current role?<br>What does an average day look like? |
| Tell me about your experience **managing heart failure patient services either in hospital or other**<br>(10 mins) | What are some of the challenges you face?<br>What are some of the challenges the health system faces?<br>What is working well? |
| What **information** helps you provide or support heart failure care?<br>(10 mins) | Where do you find the information?<br>How do you use it?<br>How is it stored?<br>**What information would be helpful that you don't have access to now?** |
| What **decisions** do you make about heart failure care?<br>(10 mins) | How do you use information to help make these decisions?<br>Are there types of information that you trust more than others? Are there types of information that have more weight than others? |
| What **tools** do you use day to day?<br>(5 mins) | Can you show me<br>What is the most useful tool or useful parts of these tools?<br>Do you use **dashboards** in your role?<br>What works well?<br>What is difficult or frustrating? |
| How could a **heart failure dashboard** support you in the future?<br>*(it could show you things like the probability of death and hospitalisation at a ward or hospital level)*<br>(10 mins) | What information would you like to see?<br>How would you use this information? What type of actions do you want to take with the information?<br>How often do you need to see the information?<br>Who else would benefit from seeing it? |
| When we are rolling out the dashboard, what should we keep front of mind?<br>(5 mins) | What are the safety considerations?<br>What are the potential barriers?<br>How would the tool fit with your workflow? |
| If you had a **magic wand** to create something that would make it easier to provide or support heart failure care, what would you make or change? | *Ask if the observer has any questions*<br>Is there anything we haven't talked about that you wish we had?<br>Any questions for me? |

### 2. Clinical dashboard stream – Clinician interview conversation guide

| Key question | Ask these as prompts if needed |
| --- | --- |
| Tell me a bit about **you and your role** | How long have you worked at _____?<br>What is your current role?<br>What does an average day look like?<br>How many heart failure patients do you usually look after over a week? |
| Talk me through the **high-level steps** you typically go through **to manage a heart failure patient** (in hospital or in the community)? | What is working well?<br>What are some of the challenges you face?<br>What are some areas for improvement?<br>How do you decide what patient to see first? |
| **Who is typically involved in the care** of a heart failure patient? (other clinicians inside and outside of hospital) | How do you work together?<br>How do you share information? |
| What **information** helps you provide or support heart failure care at the **bedside/home/in rooms**? | Where do you find the information?<br>What device do you use?<br>How often do you refer to it?<br>Do you download and store it anywhere?<br>How do you know the data is accurate, complete and up to date?<br>How easy is this information to find?<br>What information would be helpful that you don't have access to now?<br>How is information shared with patients?<br>How is information shared with other clinicians? |
| How could a patient-level heart failure dashboard **support you in the future**? | What information would you like to see?<br>How would you use this information?<br>What type of actions do you want to take with the information?<br>How often do you need to see the information?<br>Who else would benefit from seeing it?<br>How do you imagine it fitting with your workflow? |
| If you had a **magic wand** to create something that would make it easier to provide or support heart failure care, what would you make or change? | *Ask if the observer has any questions*<br>Is there anything we haven't talked about that you wish we had?<br>Any questions for me? |

### 3. Clinical Dashboard Stream – Consumer interview conversation guide

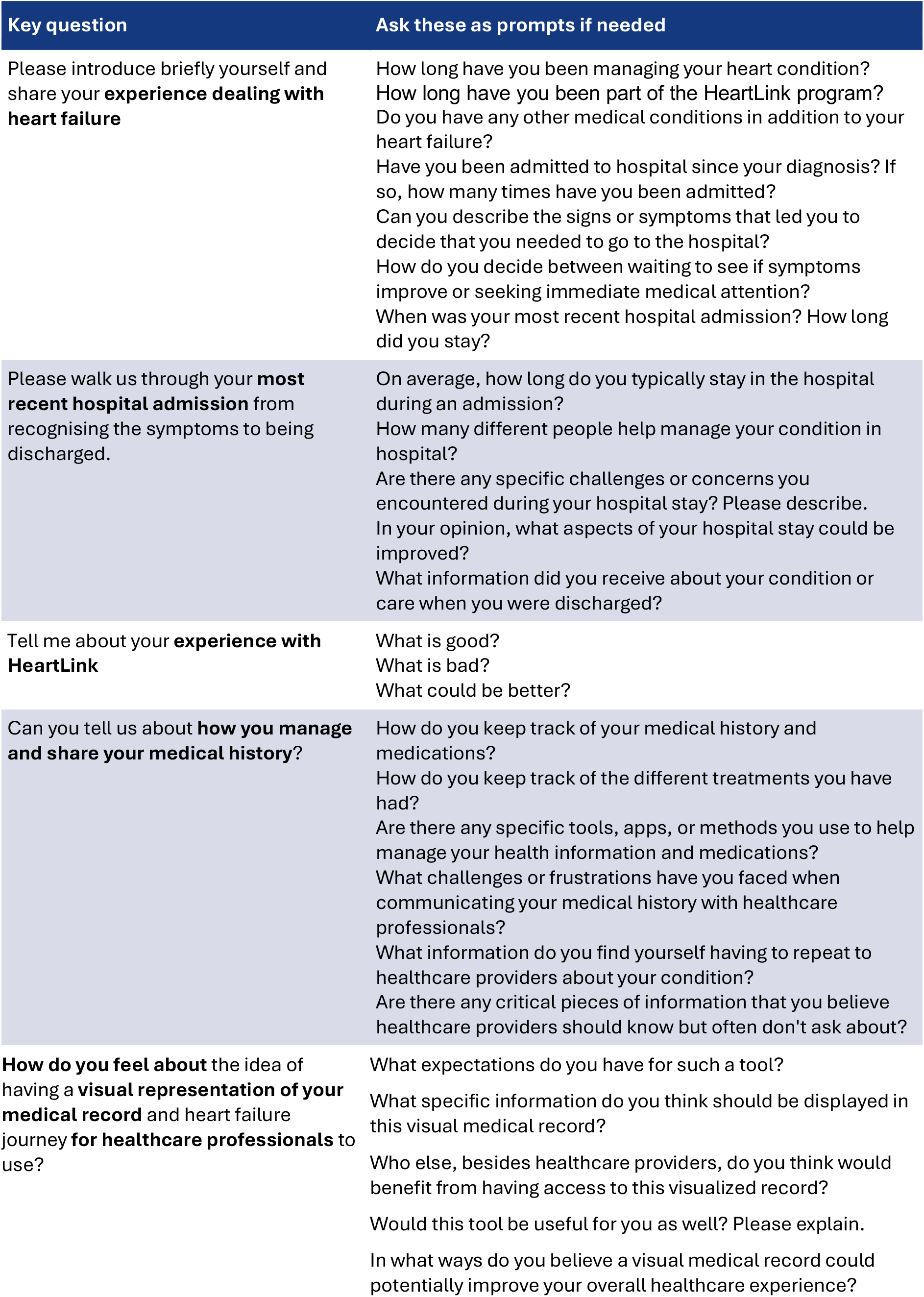
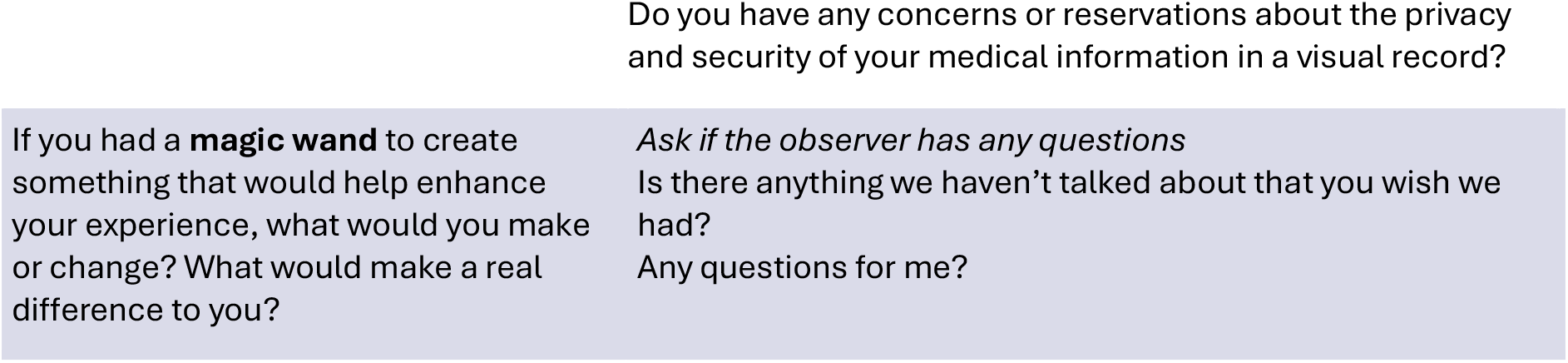

## Notes

### Competing Interest Statement

The authors have declared no competing interest.

